# Cost-effectiveness analysis of 21-valent pneumococcal conjugated vaccine among adults in Canada

**DOI:** 10.1101/2024.10.21.24315770

**Authors:** Raphael Ximenes, Alison E. Simmons, Gebremedhin B. Gebretekle, Austin Nam, Eva Wong, Marina I. Salvadori, Alyssa R. Golden, Beate Sander, Kyla J. Hildebrand, Matthew Tunis, Ashleigh R. Tuite

## Abstract

**Background:** A 21-valent pneumococcal conjugate vaccine (PCV21) was recently authorized in Canada to protect adults against invasive pneumococcal disease (IPD).

**Objective:** To assess the cost-effectiveness of PCV21 compared to current Canadian vaccination recommendations for adults of different age and risk groups.

**Methods:** We used a static cohort model to estimate lifetime incremental cost-effectiveness ratios (ICERs), in 2023 Canadian dollars per quality-adjusted life year (QALY), discounted at 1.5%, in population cohorts aged 33 (midpoint of the 18-49 year age group), 50, and 65 years from the health system and societal perspectives. The primary analysis used 2022 serotype distributions for IPD cases. Additional analyses incorporated indirect effects from pediatric vaccination and used IPD serotype distributions from 2015-2019, to explore the impact of changes over time observed in some age groups.

**Results:** For population groups currently recommended to receive PCV20 in Canada (65 years and older, 50-64 years with additional risk factors for IPD, or 18-49 years with immunocompromising conditions), PCV21 was cost-effective at a $50,000 per QALY threshold and dominated PCV20 in most scenarios when PCV21 serotypes were more prevalent. When PCV20 serotypes were equally or more prevalent than PCV21 serotypes, results were more sensitive to assumptions about indirect effects and serotype replacement. For groups not currently recommended a conjugate vaccine (50-64 years without additional IPD risk factors and 18-49 years with chronic medical conditions or unhoused populations), use of a higher-valency conjugate vaccine was a cost-effective intervention compared to no vaccination, with the optimal vaccine dependent on the proportion of IPD attributable to PCV20 and PCV21 serotypes in the population of interest. Results were sensitive to vaccine price in most scenarios.

**Interpretation:** The use of PCV21 may be cost-effective in some populations, depending on the prevalence of IPD serotypes covered by PCV20 and PCV21.

## Introduction

Invasive pneumococcal disease (IPD) is a serious and acute infectious disease with presentations such as meningitis, sepsis, and bacteremic pneumonia (1). Non-invasive forms of pneumococcal disease (PD) include community-acquired pneumonia (pCAP). PD is caused by Streptococcus pneumoniae, which has more than 100 serotypes; however, a small number of serotypes cause the majority of disease (2). The incidence of IPD in Canada peaked at 10.9 cases per 100,000 population in 2018, decreased between 2019 to 2021 due to the COVID-19 pandemic, and returned to pre-pandemic levels in 2022 (3). In Canada, PD remains a substantial public health problem, affecting most commonly young children, older adults, and individuals with specific medical conditions (e.g., chronic heart disease, chronic lung disease, chronic neurological conditions, diabetes, immunocompromising conditions) or environmental or living conditions (e.g., individuals who are unhoused, individuals who live in communities experiencing sustained high rates of IPD, individuals who use drugs) that increase PD risk (1, 4).

Vaccination is effective in reducing the incidence of PD (3). In recent years, several higher-valency pneumococcal conjugate vaccines (PCVs) have been authorized for use in Canada. At the time of this analysis, 15-valent pneumococcal conjugate vaccine (PCV15) and 20-valent pneumococcal conjugate vaccine (PCV20), which cover 15 and 20 S. pneumoniae serotypes, respectively, were recommended by Canada’s National Advisory Committee on Immunization (NACI) for use in pediatric and adult populations (5, 6). For adults recommended to receive pneumococcal vaccination, PCV20 has been preferred over PCV15 (5). A newly authorized 21-valent pneumococcal conjugate vaccine (PCV21) contains 10 unique, non-cross-reactive serotypes that are not included in PCV20 (9N, 15A, 16F, 17F, 20A, 23A, 23B, 24F, 31 and 35B). Conversely, PCV20 contains 9 unique serotypes not included in PCV21 (1, 4, 5, 6B, 9V, 14, 18C, 19F, 23F).

Vaccination program impact will be affected by the proportion of IPD cases caused by serotypes included in the vaccine. A vaccine that covers serotypes responsible for a larger share of IPD cases is expected to have a larger population impact, assuming similar vaccine effectiveness (VE) across serotypes. The distribution of vaccine-contained serotypes among IPD cases varies across adult age groups in Canada. Data from 2015–2019 and 2022 show that PCV21 generally offers broader serotype coverage of IPD cases compared to PCV20, especially in older adults (7). In adults aged 65 years and older, the serotypes contained in PCV21 accounted for substantially more IPD isolates (84.2% in 2015–2019 and 79.4% in 2022) compared to the serotypes contained in PCV20 (54.2% in 2015-2019 and 58.2% in 2022) (7). For adults aged 50–64 years, the serotypes contained in PCV21 accounted for 82.1% (2015-2019) and 70% (2022) of IPD, whereas the serotypes contained in PCV20 accounted for 62.3% (2015-2019) and 69.9% (2022). In the 18–49 year age group, PCV21-contained serotypes accounted for 76.9% (2015-2019) and 64% (2022) of IPD isolates while PCV20-contained serotypes accounted for 70.4% (2015–2019) and 78.2% (2022) of IPD isolates.

Given the different serotypes covered by PCV20 and PCV21, it is important to assess the potential impact associated with the use of these vaccines in publicly funded programs for the adult population living in Canada. In this study, we performed a model-based economic evaluation to calculate the cost-effectiveness of PCV21 compared to current vaccine recommendations. We assessed cost-utility in different population groups, based on age and risk factors, to inform vaccine recommendations.

## Methods

### Model Overview

This analysis was conducted to support updated NACI considerations on the use of pneumococcal vaccines in adults. For this analysis we modified a static Markov cohort model previously used by NACI to assess the cost-utility of PCV15 and PCV20 in adults living in Canada (8), following the Professional Society for Health Economics and Outcomes Research (ISPOR) Consolidated Health Economic Evaluation Reporting Standards 2022 (CHEERS 2022) statement (9). The model and assumptions have been described previously (8), a high-level overview is therefore provided below, with a focus on changes implemented for the current analysis. Additional details are provided in the Supplementary Materials.

The model followed a single cohort of individuals without a history of previous pneumococcal vaccination over their lifetime, using a cycle length (i.e., time step) of one-year. Vaccines were assumed to be administered at the start of the model, with vaccination lowering the risk of PD caused by the serotypes covered by the vaccine. Individuals did not have PD on model entry, but could develop IPD or pCAP over the course of their lives (Figure 1). Individuals who recovered from IPD, specifically those who had meningitis (with all cases assumed to require hospitalization), could experience long-term complications from the infection, including neurological or auditory sequelae. Baseline mortality was included, with an additional risk of death associated with PD (10). We estimated quality-adjusted life years (QALYs), costs (in 2023 Canadian dollars), and incremental cost-effectiveness ratios (ICERs) for different vaccination strategies. Lifetime costs and outcomes were discounted at 1.5% per annum and cost-effectiveness was assessed from both the health system and societal perspectives. The model was programmed using R 4.1.1 (11, 12).

**Figure 1.**
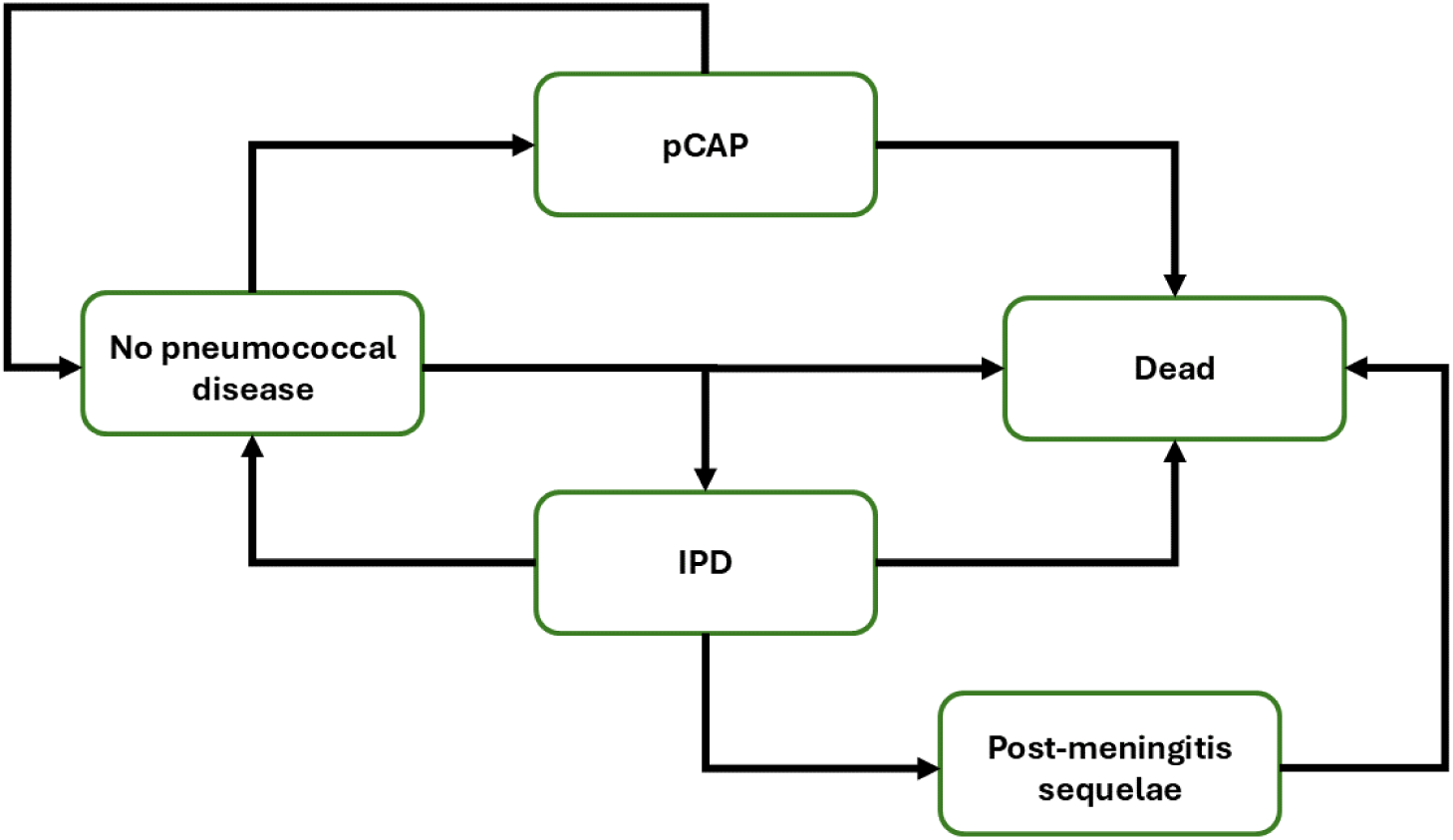
Markov model schematic of pneumococcal disease model health states. Vaccinated and unvaccinated individuals experience the same health states, with risk modified based on vaccination status, type of vaccine received, and time since vaccination. Post-meningitis sequelae includes auditory and neurologic sequelae. IPD = invasive pneumococcal disease; pCAP = non-invasive pneumococcal community-acquired pneumonia.

### Model Parameters

Model parameters and details are provided in the appendix, including those related to disease epidemiology (Table S1), vaccine characteristics (Table S2), costs (Table S3), and health state utilities (Table S4). Model inputs were obtained from available data and published studies, wherever possible, and assumptions were made when data were unavailable. Most parameters from the previously described version of the model remained unchanged (8). Key changes included updating PD incidence and serotype distribution (7), updating costs to 2023 Canadian dollars using the Consumer Price Index (13), and including parameter estimates for the population aged 18-49 years, where relevant, to represent 18-49 year old individuals with chronic medical conditions, immunocompromising conditions, and those who are unhoused.

We used data on the serotypes of IPD isolates by age to estimate the proportion of IPD and pCAP that could potentially be averted with vaccination (7). Serotypes were categorized into groups based on whether they were uniquely covered by PCV20, uniquely covered by PCV21, or shared between both vaccines (Figure 2). For vaccine serotype groupings, serotype 6C was grouped with 6A, and serotype 15C was grouped with 15B due to observed cross-protection (14, 15). Given changes in IPD epidemiology observed since the COVID-19 pandemic, we used data from the pre-pandemic period (2015-2019) as well as more recent data (for the year 2022) (Figure 2) (3). In the absence of serotyping data for pCAP, we assumed the same serotype distributions as for IPD.

**Figure 2.**
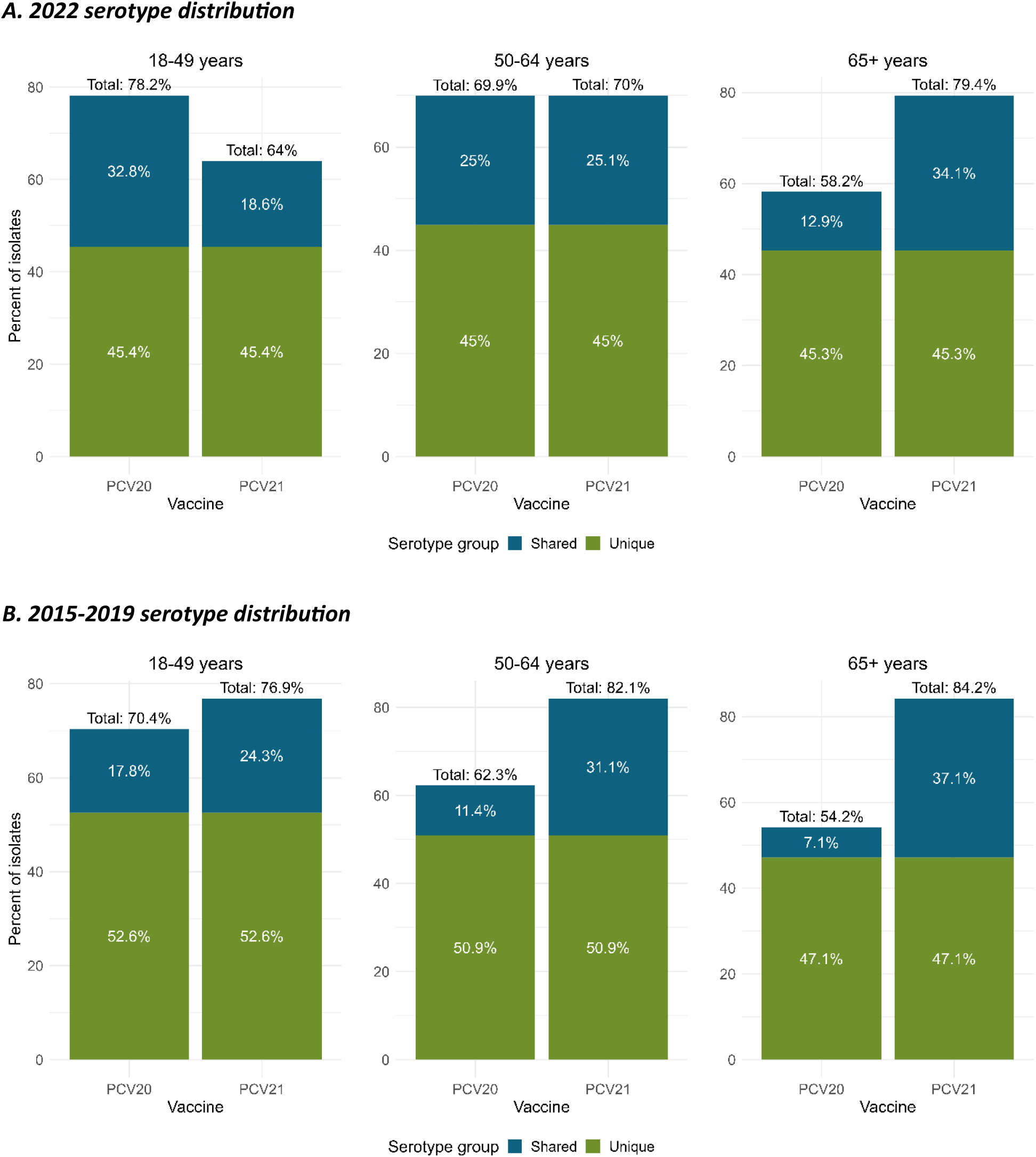
Proportion of IPD isolates caused by vaccine contained serotypes, by age group (7). A. Serotype distribution in 2022. B. Serotype distribution from 2015-2019. Graphs show the proportion of IPD isolates attributable to serotypes contained in PCV20 and PCV21. The proportion of isolates due to serotypes contained in both vaccines is shown in green and the proportion of isolates due to serotypes unique to either PCV20 or PCV21 are shown in blue.

Vaccine parameters were stratified by age and risk group (Table S2). In the absence of VE data (8), VE for PCV20 and PCV21 was assumed equivalent to VE estimates for PCV13, with coverage for the higher-valency vaccines extended to include the additional serotypes not contained in PCV13. VE for preventing PD caused by serotype 3 was assumed to be lower than for other serotypes (16, 17). Vaccination coverage data, sourced from Canadian reports (18), was incorporated to reflect current immunization rates. Duration of vaccine protection was also considered, with a 15-year horizon for conjugated vaccines, where effectiveness remains stable for 5 years, followed by a linear decline to zero over the next 10 years (19-21).

Costs included vaccination costs and PD related healthcare costs including lifetime medical costs for long-term complication. Vaccine prices were based on Canadian list prices for PCV20 ($109.91 per dose) and PCV21 ($129.90 per dose). For the societal perspective, indirect costs included productivity losses and out-of-pocket costs (Table S3).

We used age-specific utilities for the general Canadian population and for individuals with chronic medical conditions (CMC) or immunocompromising conditions (IC) (Table S4) (22). Utilities of hospitalized or outpatient PD and long-term sequelae, including auditory and neurologic complications following meningitis were derived by applying utility multipliers for each condition against the background utilities (23, 24).

### Model Population and Scenarios

We modelled single population cohorts of adults aged 33 (midpoint of 18-49 year age group), 50 (age at vaccination for the 50 – 64 year age group), and 65 years (age at vaccination for 65 years and older age group). We used age 33 years for the 18-49 year age group because we assumed that the presence of risk factors that would result in eligibility for vaccination become more common with older age and that most would not be eligible for vaccination at age 18 years. The starting age for each population cohort was selected based on current vaccine recommendations, which vary by age at vaccination. The age 33 and age 50 years cohorts were further characterized by the presence of conditions that placed individuals at increased risk of PD, based on current vaccine recommendations.

For each age and risk group, the comparator(s) were based on vaccine recommendations at the time of the analysis (5) (Table 1). Adults currently recommended a pneumococcal conjugate vaccine include: (i) individuals aged 65 years and older; (ii) individuals aged 50-64 years with additional risk factors for PD, including IC, CMC, or who are unhoused; and (iii) individuals aged 18-49 years with IC. For these population groups, we compared the use of PCV21 to the currently recommended PCV20. In a supplementary analysis, we also compared PCV21 to PPV23 at age 65 years; although PCV20 is the recommended vaccine for this group (5), PPV23 may still be considered the current standard of care in some jurisdictions while PCV20 is being integrated into vaccination programs.

**Table 1.** Vaccination strategy comparators used to evaluate the cost-effectiveness of PCV21 in different population groups.

|  | Age Group |  |  |
| --- | --- | --- | --- |
| Population Group | 18 – 49 years | 50 – 64 years | 65+ years |
| Average risk | Not evaluated | PCV20 and no vaccination | PCV20 |
| Immunocompromised (IC) | PCV20 | PCV20 | Not evaluated |
| Chronic medical conditions (CMC) | PCV20 and no vaccination | PCV20 | Not evaluated |
| Unhoused | PCV20 and no vaccination | PCV20 | Not evaluated |

Adults not currently recommended a pneumococcal conjugate vaccine who were considered in our analysis included: (i) individuals aged 50-64 years at average risk (i.e., those without additional risk factors for PD) and (ii) individuals aged 18-49 years with additional risk factors for PD, including CMC or who are unhoused. For these groups, we compared the use of PCV21 and PCV20 to no vaccination.

For the population groups who are IC, with CMC, or are unhoused, we addressed uncertainty in the magnitude of elevated risk of PD associated with these risk factors by exploring different relative risks (RR) of PD compared to the population at average risk to assess how increased risk influenced cost-effectiveness.

### Analysis

We used the estimated number of averted cases of IPD and pCAP, and associated long-term sequelae and deaths to calculate QALYs, costs, and ICERs. We conducted sequential analyses to compare all interventions considered for each population groups (25). Estimates were based on 1,000 probabilistic simulations, with parameters sampled from distributions. The number was chosen to strike a balance between computational efficiency and the precision of the results, ensuring reliable estimates without excessive computational burden. Beta distributions were used for probabilities and utilities, and gamma and lognormal distributions were used for costs and relative risks. The primary analysis used IPD isolate serotype distributions for the year 2022 without including indirect effects from pediatric vaccination or serotype replacement, given uncertainty about the magnitude of such effects. We conducted a separate analysis for Northern Canada for the age 65 years cohort and the age 50 years cohort at average risk of IPD, to explore the impact of higher PD burden and healthcare-associated costs in some Northern communities on cost-effectiveness (8). The vaccination programs’ cost-effectiveness was evaluated using a threshold of $50,000 per QALY, often used as benchmark for cost-effectiveness analyses.

We conducted one-way sensitivity analyses by varying key parameter inputs (such as vaccine price, discount rate, IPD case fatality rate and odds ratio of pCAP being treated in an outpatient setting) within plausible ranges (Table S2) while keeping all other parameters constant at their mean values. A two-way sensitivity analysis was also performed to explore the combined effect of changing both vaccine prices simultaneously (i.e., setting the maximum value for both PCV20 and PCV21 at $129.90 and the minimum at $95).

Scenario analyses included indirect effects from pediatric vaccination with PCV15 or PCV20, the vaccines currently recommended for Canadian pediatric vaccination programs, with serotype replacement. Indirect effects were modeled separately for pediatric programs using either PCV15 or PCV20. We assumed a 50% reduction in PD caused by the serotypes covered by each vaccine (excluding those in PCV13) over a period of five years, starting one year after the implementation of a pediatric vaccination program. Serotype replacement was modeled as a corresponding rise in non-PCV15 (for a PCV15 pediatric program) or non-PCV20 (for a PCV20 pediatric program) vaccine serotypes, assuming 100% replacement. We also conducted a scenario analysis using pneumococcal serotype distributions for the years 2015 to 2019, given the changes in PD epidemiology that have occurred since the COVID-19 pandemic and uncertainty about future trends. Results are summarized below for the health system perspective. Full results, including results for the societal perspective, are available in the appendix (Table S5-Table S12).

## Results

### Adults aged 65 years and older at average risk of pneumococcal disease

For adults aged 65 years and older, where the serotypes contained in PCV21 accounted for more IPD cases than PCV20 for both time periods considered, PCV21 consistently dominated PCV20 (i.e., PCV21 was more effective and less costly than PCV20) regardless of the indirect effects assumptions and serotype distribution used (Figure 3; Table S5). Similar results were observed for the Northen Canada setting, where there are higher medical costs and incidence of PD (Figure 4; Table S5). Using PPV23 as a comparator, PCV21 was dominant (i.e., more effective and less costly) (not shown). In a one-way sensitivity analysis, PCV21 remained the dominant strategy compared to PCV20 across all plausible ranges of values for parameters under consideration.

**Figure 3.**
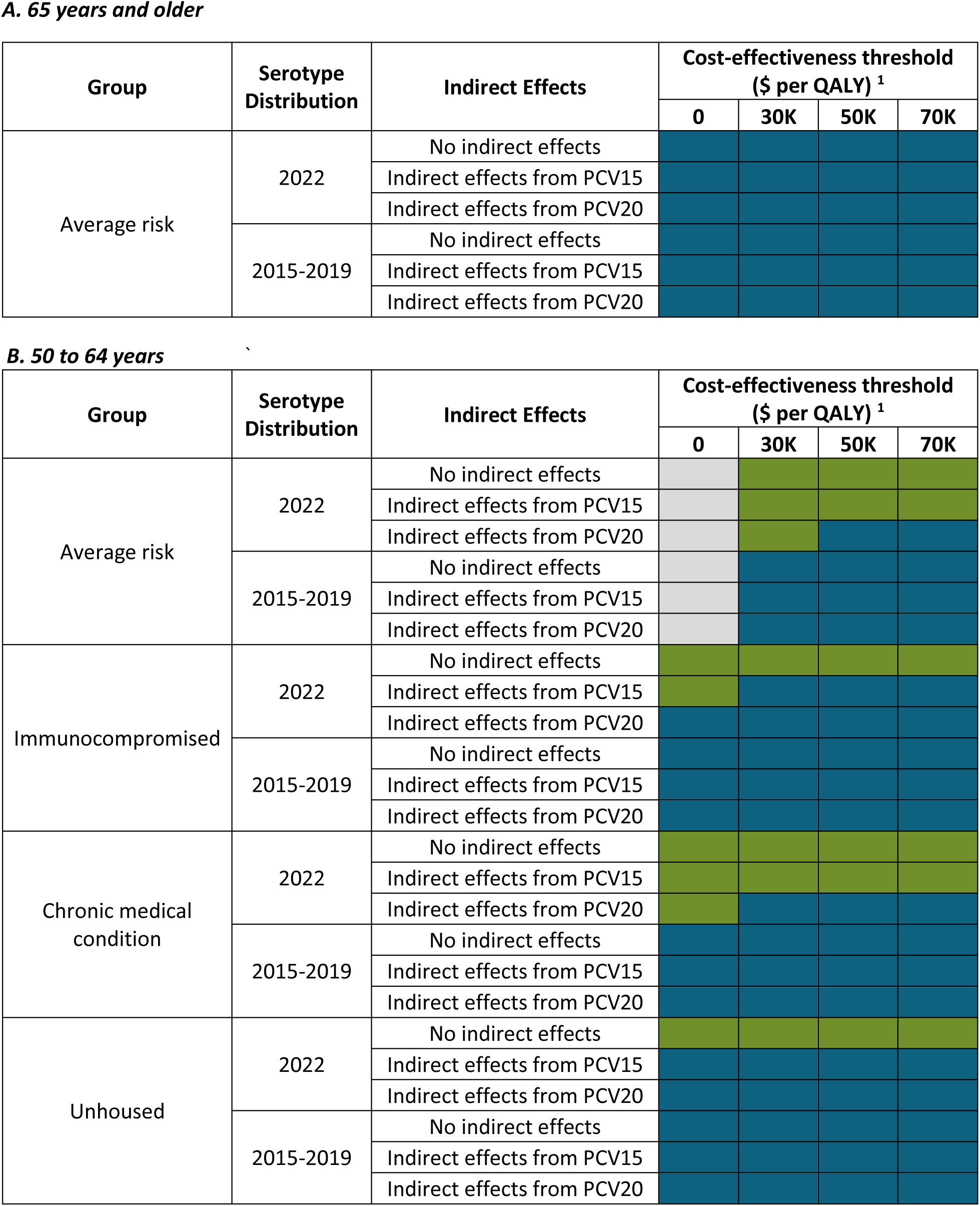
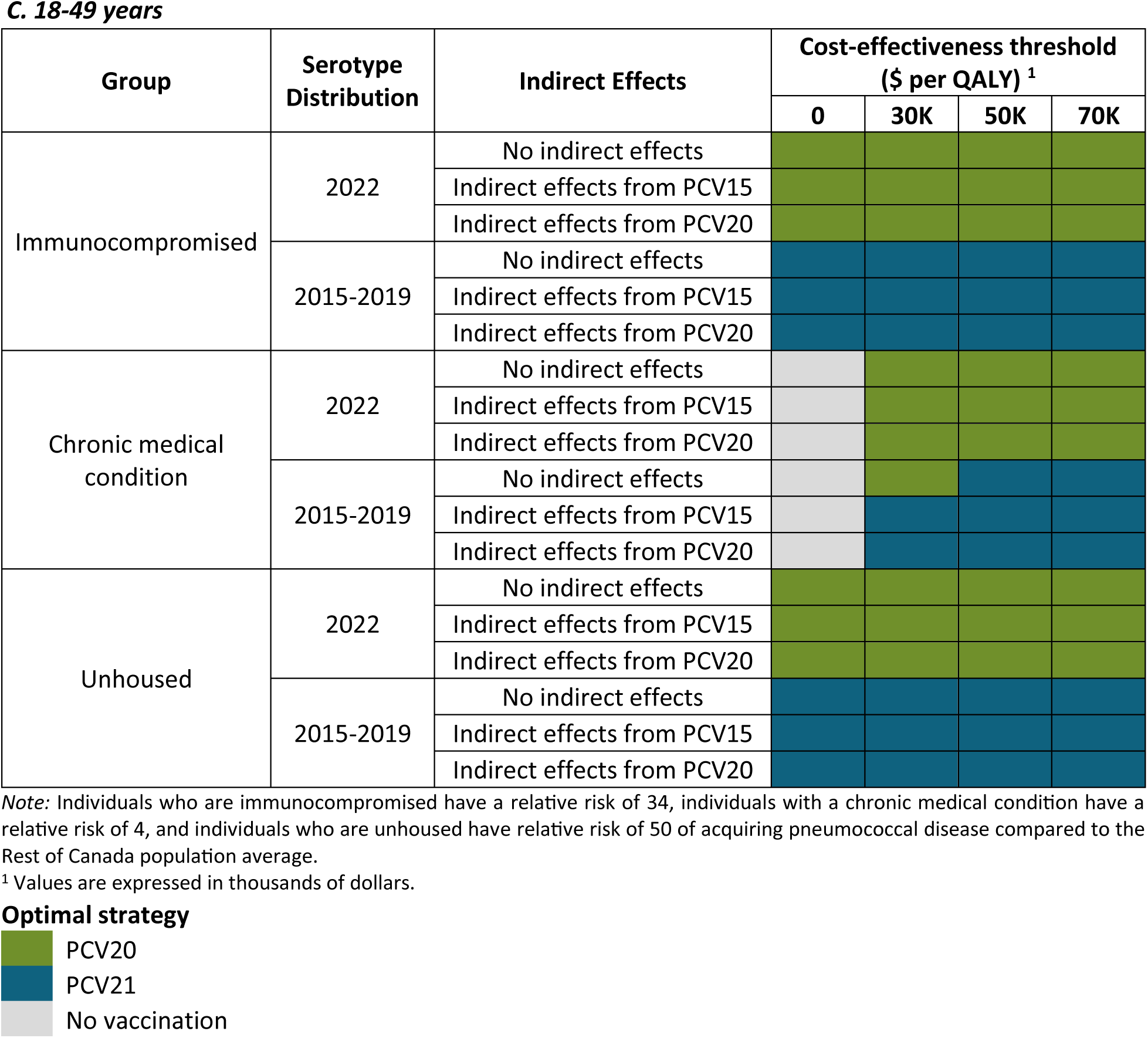
Optimal pneumococcal vaccination strategies from the health system perspective among adults aged (A) 65 years and older, (B) 50 to 64 years, and (C) 18-49 years residing in the Rest of Canada at cost-effectiveness thresholds ranging from $0/QALY gained to $70,000/QALY gained.

**Figure 4.**
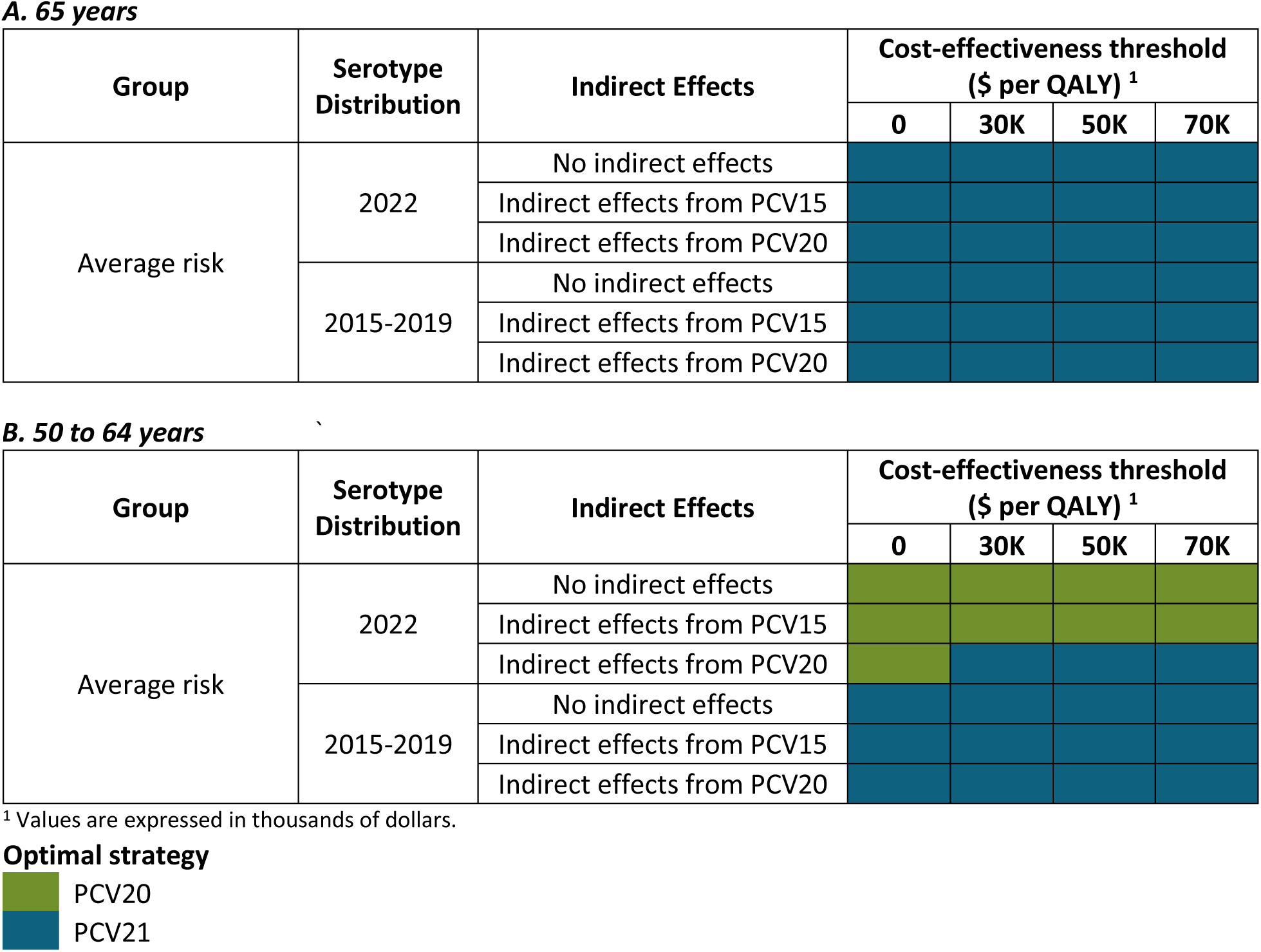
Optimal pneumococcal vaccination strategies from the health system perspective in cohorts of adults (A) 65 years and older and (B) 50 to 64 years residing in Northern Canada at cost-effectiveness thresholds ranging from $0/QALY gained to $70,000/QALY gained.

### Adults aged 50-64 years at average risk of pneumococcal disease

For adults vaccinated at age 50 years at an average risk of PD who are not currently recommended to receive a conjugate vaccine, a strategy of vaccination with one of the conjugate vaccines was optimal compared to no vaccination (Figure 3; Table S6). The optimal vaccine (i.e., PCV20 or PCV21) depended on the serotype distributions used and the assumptions of indirect effects from pediatric vaccination programs.

The use of PCV20 was cost-effective at a cost-effectiveness threshold of $50,000 per QALY when using the 2022 serotype distribution, where the prevalence of unique PCV20 and unique PCV21 serotypes were equally common, and considering no indirect effects. One-way sensitivity analysis revealed that the price of PCV21 was the only parameter influencing the optimal vaccination strategy (Table 2). The two-way sensitivity analysis indicated that a 15% reduction in the price of PCV21 would make it a cost-effective option.

**Table 2.**
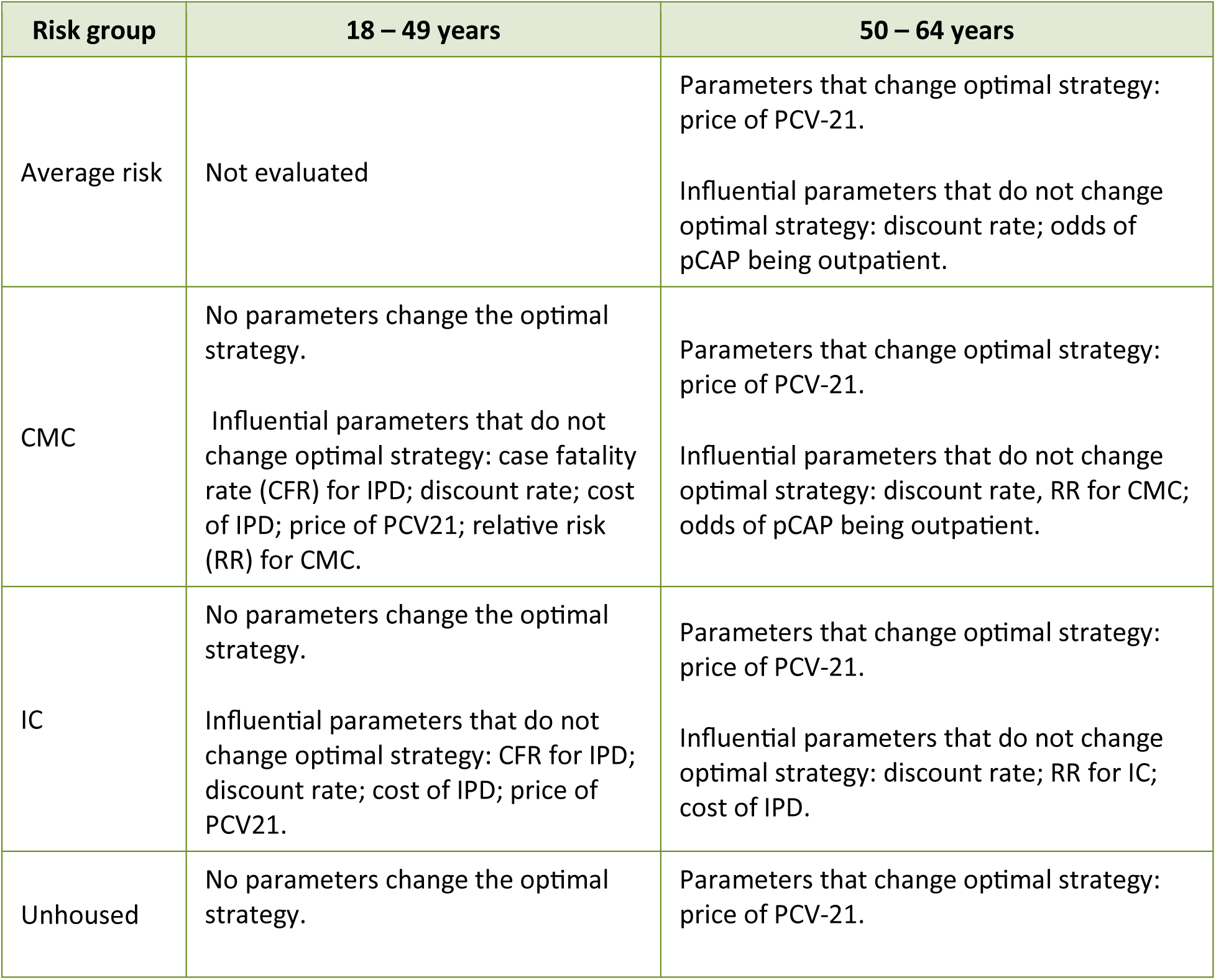
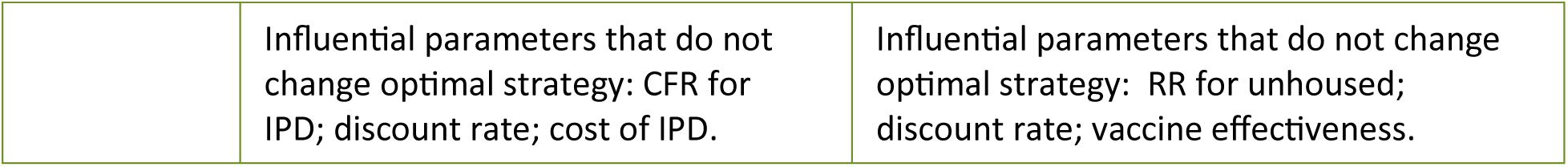
One-way sensitivity analysis of influential parameters.

Use of PCV20 remained the optimal strategy when indirect effects from PCV15 with serotype replacement were included (Figure 3; Table S6). However, PCV21 was cost-effective using the 2022 serotype distribution and assuming indirect effects from PCV20 with serotype replacement (Figure 3; Table S6). Using the serotype distribution from 2015-2019, where serotypes covered by PCV21 were more prevalent, PCV21 dominated PCV20 both with and without considering indirect effects, and was cost-effective when compared to no vaccination, with ICERs of about $15,000 per QALY.

In the Northern Canada context, using the 2022 serotype distribution, PCV20 was the optimal strategy, except when considering indirect effects from PCV20 with serotype replacement, where PCV21 was cost-effective compared to PCV20, with an ICER of $7,802 per QALY (Figure 4; Table S6). However, when the 2015-2019 serotype distribution was applied, PCV20 was dominated by PCV21 across all scenarios (Figure 4; Table S6).

### Adults aged 50-64 years with additional risk factors for pneumococcal disease

For adults vaccinated at age 50 years at high risk of IPD and currently recommended a conjugate vaccine, such as those with IC, CMC, or who are unhoused, the cost-effectiveness of PCV21 compared to PCV20 was impacted by the serotype distribution used and assumptions about indirect effects and serotype replacement due to pediatric pneumococcal vaccination programs (Figure 3; Figure 4; Table S7-Table S9).

Using the 2022 serotype distribution, where the serotypes covered by both vaccines were equal, and excluding indirect effects, PCV20 was the preferred vaccination strategy across all risk groups assessed(Figure 3; Figure 4; Table S7-Table S9). One-way sensitivity analysis showed that price of PCV21 was the only parameter influencing a change in the optimal vaccine strategy (Table 2). The two-way sensitivity analysis revealed that the price per dose of PCV21 should be reduced by at least 7% for the unhoused, 12% for IC and 15% for CMC populations to make PCV21 a cost-effective option compared to PCV20 for these population groups.

With the inclusion of indirect effects and serotype replacement, the optimal vaccine was dependent on the vaccine used in the pediatric program and the relative risk of IPD in the population group (Figure 3; Figure 4; Table S7-Table S9). PCV21 was an optimal strategy at a $50,000 per QALY threshold across all risk groups when indirect effects from PCV20 and serotype replacement were included. When indirect effects from PCV15 and serotype replacement were included, PCV21 was cost-effective for IC and unhoused populations, while PCV20 was the optimal vaccine for individuals with CMC. Using the 2015-2019 serotype distribution, where PCV21-contained serotypes were more prevalent among IPD cases, PCV21 dominated PCV20 for all scenarios (Figure 3; Figure 4; Table S7-Table S9).

### Adults aged 18-49 years with additional risk factors for IPD

For adults aged 18-49 years at high risk of IPD, specifically those with IC, CMC, or who are unhoused, the cost-effectiveness of vaccination strategies varied based on serotype distribution, inclusion of indirect effects, and the relative risk of IPD in these groups compared to average population risk. For this age group, IPD due to PCV20 serotypes were more prevalent in 2022, while IPD due to PCV21 serotypes were more common in 2015-2019.

For individuals with IC, PCV20 was the dominant vaccination strategy over PCV21 in all scenarios (Figure 3; Table S10) when using the 2022 serotype distribution. However, when the 2015–2019 serotype distribution was applied, PCV21 became the optimal choice (Figure 3; Table S10).

For both the unhoused population and the population with CMC, vaccine choice was dependent on the serotype distribution. PCV20 was the optimal vaccination strategy with 2022 serotype distribution, while PCV21 was the optimal strategy when we used the 2015-2019 serotype distribution (Figure 3; Table S11; Table S12), regardless of assumptions about indirect effects.

Varying parameters within the plausible range of values did not change the optimal vaccine choice of the primary analysis in one-way sensitivity analyses for any of these groups (Table 2).

Since unhoused individuals and individuals with CMC in this age group are not currently recommended a conjugate vaccine, we conducted an additional analysis to explore the sensitivity of our results to assumptions about the magnitude of PD risk in these groups. For unhoused individuals, we reduced the RR to 30 (compared to 50 in the primary analysis) and for individuals with CMC, we evaluated RR values of 2, 1.5 and 1.25 (compared to a RR of 4 for the primary analysis).

For the unhoused population aged 18-49 years, a strategy of vaccination with a high-valency conjugate vaccine (PCV20 or PCV21) was always identified as cost-effective compared to no vaccination, even when RR was reduced to 30, compared to 50 in the primary analysis. However, the optimal vaccine depended on the serotype distribution. Using a $50,000 per QALY cost-effectiveness threshold, PCV20 was optimal when PCV20 serotypes were more common (2022 data) and PCV21 was optimal when PCV21 serotypes were more common (2015-2019 data) (not shown).

For the population aged 18-49 years with CMCs and a $50,000 per QALY cost-effectiveness threshold, when the RR of PD was 2 (compared to 4 in the primary analysis), vaccination with PCV20 was optimal only when using the 2022 serotype distribution with no indirect effects or when indirect effects from pediatric vaccination using PCV15 and serotype replacement were included. For all other scenarios when RR was 2 (2015-2019 data and/or serotype replacement due to use of PCV20 in infants) or when RR was less than 2, a no vaccination strategy was optimal (not shown).

## Discussion

Our analysis demonstrates that PCV21 may be a cost-effective alternative to PCV20 among adults living in Canada, particularly those aged 65 years and older, where PCV21 dominated PCV20. We further found that use of a higher-valency conjugate vaccine may be cost-effective in adults aged 50 years without additional risk factors for PD and younger adults with risk factors for PD who were not currently recommended a vaccine. Our results also highlight the importance of serotype distribution for informing the optimal vaccine choice; in particular, the proportion of IPD that is covered by the serotypes in PCV20 and PCV21 was shown to be influential on estimated cost-effectiveness.

Our findings are consistent with other analyses of the cost-effectiveness of PCV21 among adults conducted in the Netherlands and the United States (26), which similarly found that PCV21 may be a cost-effective intervention in some age and risk groups (27, 28). Of note, the serotypes included in PCV21 caused more IPD cases within the population than the serotypes contained in PCV20 in the international analyses. In our analysis, we observed substantial differences in expected serotype coverage by age and time, which resulted in differences in the optimal vaccination strategy for different population groups. Our findings suggest that vaccination strategies should be tailored to current epidemiological data and may be dynamic over time and across different populations, based on trends in prevalent serotypes. In particular, indirect effects and serotype replacement following use of conjugate vaccines in pediatric populations may alter the proportion of PD that is preventable in adults by PCV20 and PCV21, potentially shifting the optimal vaccine choice as pediatric vaccination programs become established.

An important consideration is the uncertainty surrounding future pneumococcal serotype distribution. The COVID-19 pandemic disrupted patterns of pneumococcal transmission due to change in social behavior, public health measures, and healthcare practices. It remains uncertain whether the observed changes in serotype distribution will persist, revert to pre-pandemic patterns, or evolve differently. This unpredictability may impact the cost-effectiveness of PCV21 compared to PCV20, as shifts in serotype prevalence could alter the proportion of IPD cases covered by each vaccine.

This analysis is subject to limitations, including simplifying assumptions and data uncertainty. Key limitations include the assumption of equivalent VE for all serotypes in both vaccines and lack of data on serotype distribution for pCAP cases. For our analysis of risk factors aside from age, we assumed that risk of PD was elevated compared to age-specific averages, but it is important to note that there is a range of risk within the risk groups we modelled. We conducted a sensitivity analysis for the risk associated with chronic medical conditions to show how assumed risk magnitude altered our results but did not model specific risk conditions. Scenario analyses explored indirect effects and serotype replacement due to pediatric vaccination programs, but dynamic changes in disease epidemiology and vaccine impact over time were not fully captured (29). Despite these limitations, our study provides valuable insights into vaccination strategies for adults living in Canada.

Future research should consider the changes in pneumococcal serotypes causing IPD over time and across different geographic regions and any resulting effects on vaccine program impact (30). Studies that model transmission and indirect effects more comprehensively would help refine cost-effectiveness estimates. Additionally, evaluating the budget impact of vaccination strategies and exploring the cost-effectiveness of catch-up vaccination with PCV21 for those previously vaccinated with PCV20 could provide additional guidance for decision-makers.

## Conclusions

The study showed that vaccination with PCV21 is likely a cost-effective alternative to the current standard of care in Canada, particularly for adults aged 65 years. The choice between PCV21 and PCV20 is influenced by the prevailing serotype distribution, indirect effects from pediatric vaccine programs, and individual and population differences in risk, as well as cost of the vaccines. The findings provide evidence to inform policy decisions aimed at enhancing pneumococcal disease prevention in adults.

## Data Availability

All data produced in the present work are contained in the manuscript

## Authors’ statement

RX – Conceptualization, model development, data curation, analysis and interpretation of data, drafting the paper, reviewing & editing

AES – Conceptualization, data curation, analysis and interpretation of data, reviewing & editing

GBG – Conceptualization, analysis and interpretation of data, reviewing & editing

AN – Model development, Reviewing & editing.

EW – Conceptualization, Reviewing & editing.

MIS – Conceptualization, Reviewing & editing.

ARG – Conceptualization, Reviewing & editing.

BS – Conceptualization, reviewing & editing.

KJH – Conceptualization, Reviewing & editing.

MT – Conceptualization, Reviewing & editing.

ART – Conceptualization, analysis and interpretation of data, reviewing & editing.

## Acknowledgements

The authors thank members of the NACI Pneumococcal Working Group for providing feedback during model development and analysis.

## Notes

### Competing Interest Statement

The authors have declared no competing interest.

### Funding Statement

This research was supported, in part, by a Canada Research Chair in Economics of Infectious Diseases held by Beate Sander (CRC-2022-00362).

### Author Declarations

Only openly accessible public data sources or simulated data were used in this work. Details of the oversight body: All source data used in this study are available to the public and can be accessed through the references cited within the manuscript. Model parameters and additional details are provided in the Supplementary Materials. All necessary patient/participant consent (including consent to publish) has been obtained and the appropriate institutional forms have been archived and any patient/participant/sample identifiers included were not known to anyone (e.g., hospital staff, patients or participants themselves) outside the research group and cannot be used to identify individuals.

