## Supplemental files for "Cost-effectiveness analysis of 21-valent pneumococcal conjugated vaccine among adults in Canada"

Funding statement:
This research was supported, in part, by a Canada Research Chair in Economics of Infectious Diseases held by Beate Sander (CRC-2022-00362).

**Keywords:** Pneumococcal, Invasive pneumococcal disease; Vaccines, PCV20, PCV21, Cost-effectiveness, High risk populations, health economic modeling

**Appendix A. Supplementary Material**

**Supplementary Methods:** Age- and region-specific incidence data for IPD were obtained from the International Circumpolar Surveillance program and the Canadian Notifiable Disease Surveillance System (1, 2). The model accounts for regional variations, such as significantly higher incidence of IPD and CAP in Northern Canada compared to the rest of the country. The serotype distribution by age group was obtained from The Public Health Agency of Canada's National Microbiology Laboratory [PHAC-NML]) (3). Serotypes were then categorized into distinct groups based on vaccine coverage. We classified serotypes covered by multiple vaccines and identified those unique to specific vaccines.

Data on age-specific inpatient CAP incidence were obtained from the Discharge Abstract Database (DAD) (4). The relative risk of PD in higher-risk populations (5-8), as well as estimates of case fatality (9, 10) and the risk of long term-sequelae (11-13) were obtained from the literature. Duration of vaccine protection was accounted for with a 15-year time horizon for the conjugate vaccine, assuming stable effectiveness for the first 5 years, followed by a linear decline in effectiveness, reaching zero by the end of the 15-year period. (11-13).

The costs associated with IPD and pneumonia-related hospitalizations were estimated using Resource Intensity Weights obtained from the Discharge Abstract Database (4). Hospitalization costs for IPD and CAP were adjusted for different age groups and regions, taking into account the higher healthcare costs in Northern Canada (14-18). The model also considered non-medical costs, such as transportation, which can be substantial in remote areas (4, 19-27). Additionally, indirect costs, such as lost productivity and caregiver burden (28-30), were included in the economic analysis for the societal perspective.

***Table S1***. *Epidemiologic parameters*

| Parameter | Base | Range | Reference |
|---|---|---|---|
| **IPD incidence (per 100,000)** | | | |
| **18-49 years** | | | Canadian Notifiable Disease System (CNDSS) 2018-2019; International Circumpolar Surveillance (ICS) program 2018-2019 (1, 2) |
| Northern Canada[a] | 11.2 | 8.4 – 14.0[*] | |
| Rest of Canada | 5.2 | 3.9 – 6.5[*] | |
| **50-64 years** | | | |
| Northern Canada | 38.97 | 23.10 – 58.93 | |
| Rest of Canada | 14.45 | 13.83 – 15.09 | |
| **65-74 years** | | | |
| Northern Canada | 71.30 | 34.20 – 121.79 | |
| Rest of Canada | 20.61 | 19.52 – 21.72 | |
| **75+ years** | | | |
| Northern Canada | 105.01 | 38.55 – 204.12 | |
| Rest of Canada | 31.06 | 29.51 – 32.65 | |
| **Inpatient CAP incidence (per 100,000)** | | | |
| **18-49 years** | | | DAD 2018-2019 (4) |
| Northern Canada | 167.37 | 143.86 – 190.88 | |
| Rest of Canada | 90.60 | 89.42 – 91.78 | |
| **50-64 years** | | | |
| Northern Canada | 568.81 | 500.44 – 637.18 | |
| Rest of Canada | 347.81 | 344.45 – 351.16 | |
| **65-74 years** | | | |
| Northern Canada | 1,777.32 | 1,562.12 – 1,992.51 | |
| Rest of Canada | 871.48 | 863.79 – 879.18 | |
| **75+ years** | | | |
| Northern Canada | 5,104.13 | 4,540.99 – 5,667.28 | |
| Rest of Canada | 2,845.89 | 2,829.82 – 2,861.95 | |
| **Relative risk of PD in higher risk populations** | | | |
| Experiencing homelessness | 50 | 27 – 72 | Tyrell et al., 2021; Plevnesi et al., 2009; Shariatzadeh et al., 2005; Steinberg et al., 2023 (31-34) |
| **18-64 years** | | | |
| Chronic medical condition | 4 | 3 – 6 | Shigayeva et al., 2022 (5) |
| Immunocompromising condition | 34 | 21 – 55 | |
| **65+ years** | | | |
| Chronic medical condition | 1.7 | 1.0 – 2.5 | Shigayeva et al., 2022 (5) |
| Immunocompromising condition | 4.3 | 2.5 – 7.4 | |
| **Proportion of CAP cases managed in inpatient setting (%)[c]** | | | |
| 18-49 years | 3.27 | 0.69 – 5.86 | O'Reilly et al., 2023 (14) |
| 50-64 years | 15.10 | 12.80 – 17.40 | |
| 65-74 years | 24.76 | 22.66 –26.85 | |
| 75+ years | 32.65 | 30.53 – 34.76 | |
| **Proportion of CAP cases attributed to *S. pneumoniae* (%)[d]** | | | |
| 18-49 years | 19.5 | 14.6 – 24.4[*] | LeBlanc et al., 2022 (6) |
| 50+ years | 18.0 | 8.0 – 24.0 | Lansbury et al., 2022; LeBlanc et al., 2022 (6, 35) |
| **Proportion of IPD cases with meningitis (%)** | | | |

| | | | |
|---|---|---|---|
| 18-64 years | 7.0 | 5.3 – 8.8* | Backhaus et al., 2016 (9) |
| 65+ years | 5.0 | 3.8 – 6.3* | |
| **Proportion of IPD cases with meningitis who experience post-meningitis sequelae (%)** | | | |
| Neurologic sequelae[e] | 12.2 | 5.3 – 19.1 | Jit, 2010 (10) |
| Auditory sequelae[f] | 20.9 | 17.1 – 24.7 | |
| **Case fatality (%)** | | | |
| **IPD** | | | Wijayasri et al., 2019 (7) |
| 18-49 years | 5.7 | 4.9 – 6.6 | |
| 50-64 years | 10.9 | 9.9 – 12.0 | |
| 65+ years | 17.2 | 16.2 – 18.3 | |
| **Inpatient pCAP** | | | LeBlanc et al., 2022 (6) |
| 16-49 years | 3.8 | 1.4 – 6.2 | |
| 50-64 years | 4.8 | 2.7 – 6.9 | |
| 65+ years | 9.9 | 7.5 – 12.2 | |
| **Relative risk of death from PD in higher risk population** | | | |
| Chronic medical condition | 1.5 | | Shigayeva et al., 2022 (5) |
| Immunocompromising condition | 1.9 | | Shigayeva et al., 2016 (8) |

*Notes:* IPD = invasive pneumococcal disease; CAP = community acquired pneumonia.

[a] Values for Northern Canada were calculated by adjusting estimates for the Rest of Canada with multipliers (i.e., 2.15) extracted from the Canadian Notifiable Disease System (CNDSS) 2018-2019 and International Circumpolar Surveillance (ICS) program 2018-2019 (1, 2)

[b] Parameters were derived using polynomial regression.

[d] Proportion of CAP cases among hospitalized adults where *S. pneumoniae* was detected from blood culture, sputum culture, or urine antigen detection

[e] Cranial nerve palsies

[f] Hearing loss

* Range defined as ±25% of the base value

***Table S2****. Vaccine parameters*

| Parameter | Base | Range | Reference |
|---|---|---|---|
| **Vaccination coverage (%)** | | | |
| 18-49 years | 21.2 | 19.1 – 23.5 | Government of Canada, 2024 (36) |
| 50-64 years | 28.8 | 26.5 – 31.1 | |
| 65-79 years | 54.0 | 51.9 – 56.0 | |
| **With chronic medical conditions (including immunocompromising conditions)** | | | |
| 18-49 years | 24.3 | 21.6 – 27.3 | Government of Canada, 2024 (36) |
| 50-64 years | 33.4 | 30.8 – 36.2 | |
| 65-79 years | 60.7 | 57.7 – 63.7 | |
| 80+ years | 65.6 | 56.1 –74.0 | |
| **Vaccine effectiveness** | | | |
| **Pneu-C at age 65 (%)** | | | |
| VT-IPD | 88 | 61 – 98 | van Werkhoven et al., 2015 (12) |
| ST3-IPD | 53 | 0 – 80 | Lewis et al., 2019; Farrar et al., 2023 (37, 38) |
| VT-pCAP | 68 | 39 – 85 | van Werkhoven et al., 2015 (12) |
| ST3-pCAP | 16 | 0 – 32 | Stoecker, 2024 (39) |
| **Pneu-P at age 65 (%)** | | | |
| VT-IPD | 52 | 36 – 64 | Farrar et al., 2023 (37) |
| **ST3-IPD** | 2 | 0 – 21 | Djennad et al., 2018 (40) |
| **VT-pCAP** | 26 | 0 – 49 | Farrar et al., 2023 (37) |
| ST3-pCAP | 2 | 0 – 21 | Assumption |
| **Pneu-C in adults with chronic medical conditions (%)** | | | |
| VT-IPD | 75 | 41 – 98 | Bonten et al., 2014; van Werkhoven et al., 2015; Stoecker, 2024 (12, 41, 42) |
| ST3-IPD | 26 | 0 – 53 | Stoecker, 2024 (39) |
| VT-pCAP | 40 | 11 – 60 | Suaya et al., 2018 (43) |
| ST3-pCAP | 16 | 0 – 32 | Stoecker, 2024 (39) |
| **Pneu-C in adults with immunocompromising conditions (%)** | | | |
| VT-IPD | 67 | 0 – 99 | Bonten et al., 2014 (41) |
| ST3-IPD | 9 | 0 – 18 | Stoecker, 2024 (39) |
| VT-pCAP | 15 | 5 - 22 | Stoecker, 2024 (39) |
| ST3-pCAP | 5 | 0 – 11 | Stoecker, 2024 (39) |
| **Duration of protection** | | | |
| Pneu-C | 15 years: stable for 5 years, linear decline to 0 over 10 years | | Leidner, 2021; Patterson et al., 2016; van Werkhoven et al., 2015 (11-13) |
| Pneu-P | 15 years: linear decline to 0 over 15 years | | Leidner, 2021 (11) |

*Notes:* Pneu-C = pneumococcal conjugate vaccine; Pneu-P = pneumococcal polysaccharide vaccine; VT = vaccine-type serotypes; ST3 = serotype 3.

***Table S3****. Direct and indirect cost parameters*

| Parameter | Base | Range | Reference |
|---|---|---|---|
| **Cost per dose of vaccine ($)** | | | |
| Vaccine administration | 18 | 13 – 22 | O'Reilly et al., 2017 (44) |
| PCV-20 | 109.91 | | Pfizer (45) |
| PCV-21 | 129.90 | | Merck (46) |
| PPV-23 | 35.24 | | Merck (46) |
| **Cost per inpatient IPD case ($)** | | | |
| 18-49 years | 30,440 | 28,059 – 32,915 | DAD 2015-2019 (15-18) |
| 50-64 years | 30,793 | 28,909 – 32,735 | |
| 65-74 years | 30,590 | 28,237 – 33,038 | |
| 75+ years | 22,716 | 21,131 – 24,357 | |
| **Cost per inpatient CAP case ($)** | | | |
| **18-64 years** | | | O'Reilly et al., 2023 (14) |
| Northern Canada[a] | 17,906 | 17,305 – 18,538 | |
| Rest of Canada | 14,986 | 14,483 – 15,515 | |
| **65+ years** | | | |
| Northern Canada[a] | 15,436 | 15,167 – 15,713 | |
| Rest of Canada | 14,980 | 14,718 – 15,248 | |
| **Cost per outpatient CAP case ($)** | | | |
| 18-64 years | 1,255 | 1,219 – 1,290 | O'Reilly et al., 2023 (14) |
| 65+ years | 3,531 | 3,469 – 3,592 | |
| **Cost of care for patients with post-meningitis sequelae per year ($)** | | | |
| Annual cost of care for those with auditory sequelae[b] | 3,183 | 2,387 – 3,979[*] | Christensen et al., 2014 (47) |
| Annual cost of care for those with neurologic sequelae[c] | 10,593 | 7,945 – 13,241[*] | |
| **Cost of medication, out-of-pocket ($)** | | | |
| 18-64 years | 19.08 | 14.32 – 28.86 | Ontario Drug Benefit (48) |
| 65+ years | 0 | | |
| **Cost of transportation, health system ($)** | | | |
| **Inpatient** | | | |
| Northern Canada | 8,001 | 3,171 – 12,887 | DAD 2018-2019; Glauser et al., 2015; Government of Northwest Territories 2018; Government of Nunavut; Rendell, 2016; Tam et al., 2009 (4, 19-23) |
| Rest of Canada | 418 | 210 – 626 | DAD 2018-2019; Glauser et al., 2015; Government of Nunavut; Rendell, 2016 (4, 19, 21, 22) |
| **Outpatient** | | | |
| Northern Canada | 129 | 97 – 161 | Government of Yukon; Pong and Pitblado, 2005 (24, 25) |
| Rest of Canada | 0 | | Assumed to be out-of-pocket |
| **Cost of transportation, out-of-pocket ($)** | | | |
| **Inpatient** | | | |
| Northern Canada | 153 | 75 – 405 | DAD 2018-2019; Glauser et al., 2015; Government of Nunavut; Rendell, 2016 (4, 19, 21, 22) |

| | | | |
|---|---|---|---|
| Rest of Canada | 89 | 53 – 128 | DAD 2018-2019; CRA 2022; Colbert 2020 (4, 26, 27) |
| **Outpatient** | | | |
| Northern Canada | 149 | 112 – 186* | Government of Yukon; Pong and Pitblado, 2005; CRA 2022 (24-26) |
| Rest of Canada | 4 | 3 – 5* | Pong and Pitblado, 2005; CRA 2022 (25, 26) |
| Parking | 16 | 0 – 32 | Assumption |
| **Daily cost of travel subsidy for overnight stay, health system ($)** | | | |
| Northern Canada | 164 | 82 – 327 | Government of Yukon (24) |
| Rest of Canada | 0 | | |
| **Mean annual income ($)** | | | |
| 18-49 years[d] | 57,423 | | Statistics Canada (49) |
| 50-64 years[e] | 80,810 | | |
| 65+ years | 50,741 | | |
| **Mean hourly caregiver income ($)** | | | |
| All ages | 28.67 | 24.64 – 37.89 | Hollander et al., 2009 (28) |
| **Work time lost (days)** | | | |
| Inpatient IPD or CAP | 14.6 | 8.8 – 28.2 | Pasquale et al. 2019 (50) |
| Outpatient CAP | 5.4 | 1.8 – 6.3 | |
| Visit health care provider for vaccination | 0.5 | | Assumption |
| **Reduction in employment due to post-meningitis sequelae (%)** | | | |
| Auditory sequelae | 25 | 15 – 35 | Assumption used for lower estimate; Bizier et al., 2016; Jiang et al., 2012 (51, 52) |
| Neurologic sequelae | 98 | 75 – 100 | Assumption; Jiang et al., 2012 (52) |
| **Caregiver time lost (hours)** | | | |
| Inpatient IPD or CAP | 40.65 | 11.63 – 81.30 | Wyrwich et al., 2015 (29) |
| Neurologic sequelae | 5,660 | 4,245 – 7,075* | Ganapathy et al., 2015 (30) |

*Notes:* Costs were converted to Canadian dollars using purchasing power parities from the Organisation for Economic Co-operation and Development and inflated to 2023 dollars using the Canadian Consumer. Price Index. Product-specific values were used for health-related costs and transportation costs.

[a] Values for Northern Canada were calculated by adjusting estimates for the Rest of Canada with multipliers extracted from the Discharge Abstract Database 2015-2019 (15-18)

[b] The annual cost for auditory sequelae reflects the annual cost The shifting epidemiology and serotype distribution of invasive pneumococcal disease in Ontario, Canada, 2007-2017of minor post-meningitis sequelae.

[c] The annual cost of neurologic sequelae reflects the annual cost of severe post-meningitis sequelae.

[d] Median total income in 2022 CAD, inflated to 2023 CAD, for 25 to 34 years of age

[e] Median total income in 2022 CAD, inflated to 2023 CAD, for 45 to 54 years of age

* Range defined as ±25% of the base value

***Table S4**. Health utilities*

| Parameter | Base | Range | Reference |
|---|---|---|---|
| **Background health utility** | | | |
| 18-49 years | 0.873 | 0.857 – 0.89 | Yan et al., 2023 (53) |
| 50-64 years | 0.847 | 0.83 – 0.864 | |
| 65-74 years | 0.867 | 0.849 – 0.885 | |
| 75+ years | 0.861 | 0.835 – 0.887 | |
| **Average background health utility** | | | |
| Chronic medical or immunocompromising condition[a] | 0.836 | | |
| No chronic medical or immunocompromising condition | 0.899 | | |
| **IPD and pCAP utility multipliers** | | | |
| Hospitalization | 0.8659 | 0.8323 – 0.8963 | Mangen et al., 2017 (54) |
| Outpatient CAP | 0.9938 | 0.9917 – 0.9956 | Oppong et al., 2013a; Oppong et al., 2013b (55, 56) |
| Auditory Sequelae | 0.6850 | 0.6214 – 0.7451 | Galante et al., 2011 (57) |
| Neurologic Sequelae | 0.3441 | 0.2725 – 0.4164 | Galante et al., 2011 (57) |

[a] We assumed this background health utility also applies to individuals experiencing homelessness

***Table S1***. *Mean Costs, QALYs, and ICERs for pneumococcal vaccination of adults aged 65 years and older at average risk of IPD, from health system and societal perspectives (per 100,000 population)*

| Serotype distribution data year | Indirect effects assumption | Rest of Canada | | | | Northern Canada | | | |
|---|---|---|---|---|---|---|---|---|---|
| | | Strategies | Costs ($) | QALYs | Sequential ICER | Strategies | Costs ($) | QALYs | Sequential ICER |
| **Health system perspective** | | | | | | | | | |
| 2022 | No indirect Effects | PCV21 | 127,406,265 | 1,378,631 | --- | PCV21 | 260,578,720 | 1,173,753 | --- |
| | | PCV20 | 129,647,781 | 1,378,472 | Dominated | PCV20 | 268,584,936 | 1,173,461 | Dominated |
| | Indirect Effects from PCV15 | PCV21 | 127,564,622 | 1,378,624 | --- | PCV21 | 260,999,450 | 1,173,741 | --- |
| | | PCV20 | 130,121,939 | 1,378,451 | Dominated | PCV20 | 269,842,046 | 1,173,423 | Dominated |
| | Indirect Effects from PCV20 | PCV21 | 127,821,235 | 1,378,613 | --- | PCV21 | 261,681,134 | 1,173,720 | --- |
| | | PCV20 | 130,890,091 | 1,378,416 | Dominated | PCV20 | 271,877,732 | 1,173,362 | Dominated |
| 2015-2019 | No indirect Effects | PCV21 | 126,356,529 | 1,378,682 | --- | PCV21 | 257,733,767 | 1,173,844 | --- |
| | | PCV20 | 129,975,821 | 1,378,456 | Dominated | PCV20 | 269,522,644 | 1,173,430 | Dominated |
| | Indirect Effects from PCV15 | PCV21 | 126,536,703 | 1,378,673 | --- | PCV21 | 258,212,761 | 1,173,829 | --- |
| | | PCV20 | 130,514,889 | 1,378,432 | Dominated | PCV20 | 270,951,520 | 1,173,387 | Dominated |
| | Indirect Effects from PCV20 | PCV21 | 126,808,856 | 1,378,661 | --- | PCV21 | 258,936,168 | 1,173,808 | --- |
| | | PCV20 | 131,328,910 | 1,378,395 | Dominated | PCV20 | 273,108,195 | 1,173,322 | Dominated |
| **Societal perspective** | | | | | | | | | |
| 2022 | No indirect Effects | PCV21 | 164,786,256 | 1,378,631 | --- | PCV21 | 321,415,627 | 1,173,753 | --- |
| | | PCV20 | 168,082,639 | 1,378,472 | Dominated | PCV20 | 331,911,406 | 1,173,461 | Dominated |
| | Indirect Effects from PCV15 | PCV21 | 164,993,185 | 1,378,624 | --- | PCV21 | 321,946,568 | 1,173,741 | --- |
| | | PCV20 | 168,702,229 | 1,378,451 | Dominated | PCV20 | 333,497,831 | 1,173,423 | Dominated |
| | Indirect Effects from PCV20 | PCV21 | 165,328,503 | 1,378,613 | --- | PCV21 | 322,806,819 | 1,173,720 | --- |
| | | PCV20 | 169,705,986 | 1,378,416 | Dominated | PCV20 | 336,066,804 | 1,173,362 | Dominated |
| 2015-2019 | No indirect Effects | PCV21 | 163,414,965 | 1,378,682 | --- | PCV21 | 317,823,008 | 1,173,844 | --- |
| | | PCV20 | 168,528,715 | 1,378,456 | Dominated | PCV20 | 333,139,604 | 1,173,430 | Dominated |
| | Indirect Effects from PCV15 | PCV21 | 163,650,399 | 1,378,673 | --- | PCV21 | 318,427,470 | 1,173,829 | --- |
| | | PCV20 | 169,233,122 | 1,378,432 | Dominated | PCV20 | 334,942,787 | 1,173,387 | Dominated |
| | Indirect Effects from PCV20 | PCV21 | 164,006,022 | 1,378,661 | --- | PCV21 | 319,340,367 | 1,173,808 | --- |
| | | PCV20 | 170,296,818 | 1,378,395 | Dominated | PCV20 | 337,664,437 | 1,173,322 | Dominated |

***Table S2**. Mean Costs, QALYs, and ICERs for pneumococcal vaccination of adults aged 50 years at average risk of IPD, from health system and societal perspectives (per 100,000 population)*

| Serotype distribution data year | Indirect effects assumption | Rest of Canada | | | | Northern Canada | | | |
|---|---|---|---|---|---|---|---|---|---|
| | | Strategies | Costs ($) | QALYs | Sequential ICER | Strategies | Costs ($) | QALYs | Sequential ICER |
| **Health system perspective** | | | | | | | | | |
| 2022 | No indirect Effects | No vaccine | 121,709,132 | 2,099,334 | --- | PCV20 | 266,005,115 | 1,892,721 | --- |
| | | PCV20 | 123,337,835 | 2,099,437 | 15,840 | PCV21 | 266,563,217 | 1,892,721 | 1,765,679 |
| | | PCV21 | 123,901,032 | 2,099,437 | 3,246,243 | No vaccine | 267,888,430 | 1,892,530 | Dominated by PCV20 |
| | Indirect Effects from PCV15 | No vaccine | 121,709,132 | 2,099,334 | --- | PCV20 | 266,138,328 | 1,892,717 | --- |
| | | PCV20 | 123,386,599 | 2,099,434 | 16,704 | PCV21 | 266,607,624 | 1,892,720 | 145,953 |
| | | PCV21 | 123,917,288 | 2,099,436 | 299,422 | No vaccine | 267,888,430 | 1,892,530 | Dominated by PCV20 |
| | Indirect Effects from PCV20 | No vaccine | 121,709,132 | 2,099,334 | --- | PCV20 | 266,669,455 | 1,892,699 | --- |
| | | PCV20 | 123,581,051 | 2,099,425 | 20,604 | PCV21 | 266,784,721 | 1,892,714 | 7,802 |
| | | PCV21 | 123,982,116 | 2,099,433 | 49,201 | No vaccine | 267,888,430 | 1,892,530 | Dominated by PCV20 |
| 2015-2019 | No indirect Effects | No vaccine | 121,709,132 | 2,099,334 | --- | PCV21 | 265,299,501 | 1,892,764 | --- |
| | | PCV21 | 123,437,274 | 2,099,460 | 13,662 | No Vaccine | 267,888,430 | 1,892,530 | Dominated by PCV20 |
| | | PCV20 | 123,526,358 | 2,099,427 | Dominated by PCV21 | PCV20 | 266,530,502 | 1,892,702 | Dominated by PCV21 |
| | Indirect Effects from PCV15 | No vaccine | 121,709,132 | 2,099,334 | --- | PCV21 | 265,388,371 | 1,892,761 | --- |
| | | PCV21 | 123,469,789 | 2,099,459 | 14,098 | No Vaccine | 267,888,430 | 1,892,530 | Dominated by PCV20 |
| | | PCV20 | 123,623,802 | 2,099,422 | Dominated by PCV21 | PCV20 | 266,796638 | 1,892,694 | Dominated by PCV21 |

| | | | | | | | | | |
|---|---|---|---|---|---|---|---|---|---|
| | Indirect Effects from PCV20 | No vaccine | 121,709,132 | 2,099,334 | --- | PCV21 | 265,572,690 | 1,892,755 | --- |
| | | PCV21 | 123,537,230 | 2,099,455 | 15,037 | No Vaccine | 267,888,430 | 1,892,530 | Dominated by PCV20 |
| | | PCV20 | 123,825,880 | 2,099,412 | Dominated by PCV21 | PCV20 | 267,348,446 | 1,892,676 | Dominated by PCV21 |
| **Societal perspective** | | | | | | | | | |
| 2022 | No indirect Effects | No vaccine | 161,968,709 | 2,099,334 | --- | No vaccine | 343,080,452 | 1,892,530 | --- |
| | | PCV20 | 173,493,328 | 2,099,437 | 112,084 | PCV20 | 349,569,706 | 1,892,721 | 34,016 |
| | | PCV21 | 174,054,200 | 2,099,437 | 3,250,765 | PCV21 | 350,122,989 | 1,892,721 | 1,750,430 |
| | Indirect Effects from PCV15 | No vaccine | 161,968,703 | 2,099,334 | --- | No vaccine | 343,080,452 | 1,892,530 | --- |
| | | PCV20 | 173,574,903 | 2,099,434 | 115,575 | PCV20 | 349,770,210 | 1,892,717 | 35,885 |
| | | PCV21 | 174,081,392 | 2,099,436 | 285,769 | PCV21 | 350,189,827 | 1,892,720 | 130,503 |
| | Indirect Effects from PCV20 | No vaccine | 161,968,703 | 2,099,334 | | No vaccine | 343,080,452 | 1,892,530 | --- |
| | | PCV21 | 174,189,842 | 2,099,433 | 123,442 | PCV21 | 350,456,380 | 1,892,714 | 40,118 |
| | | PCV20 | 173,900,196 | 2,099,425 | Extendedly dominated | PCV20 | 350,569,624 | 1,892,699 | Dominated by PCV21 |
| 2015-2019 | No indirect Effects | No vaccine | 161,968,703 | 2,099,334 | --- | No vaccine | 343,080,452 | 1,892,530 | --- |
| | | PCV21 | 173,275,973 | 2,099,460 | 89,392 | PCV21 | 348,213,134 | 1,892,764 | 21,971 |
| | | PCV20 | 173,816,232 | 2,099,427 | Dominated by PCV21 | PCV20 | 350,379,417 | 1,894,702 | Dominated by PCV21 |
| | Indirect Effects from PCV15 | No vaccine | 161,968,703 | 2,099,334 | --- | No vaccine | 343,080,452 | 1,892,530 | --- |
| | | PCV21 | 173,330,365 | 2,099,459 | 90,973 | PCV21 | 348,346,893 | 1,892,761 | 22,827 |
| | | PCV20 | 173,979,242 | 2,099,422 | Dominated by PCV21 | PCV20 | 350,779,983 | 1,892,694 | Dominated by PCV21 |
| | Indirect Effects from PCV20 | No vaccine | 161,968,703 | 2,099,334 | --- | No vaccine | 343,080,452 | 1,892,530 | --- |
| | | PCV21 | 173,443,184 | 2,099,455 | 94,385 | PCV21 | 348,624,316 | 1,892,755 | 24,673 |
| | | PCV20 | 174,317,292 | 2,099,412 | Dominated by PCV21 | PCV20 | 351,610,525 | 1,892,676 | Dominated by PCV21 |

***Table S3**. Mean Costs, QALYs, and ICERs for pneumococcal vaccination of adults aged 50 years with immunocompromising conditions (RR = 34), from health system and societal perspectives (per 100,000 population)*

| **Serotype distribution data year** | **Indirect effects assumption** | **Rest of Canada** | | | |
|---|---|---|---|---|---|
| | | **Strategies** | **Costs** | **QALYs** | **Sequential ICER** |
| **Health system perspective** | | | | | |
| 2022 | No indirect Effects | PCV20 | 2,257,012,222 | 1,691,426 | |
| | | PCV21 | 2,257,633,083 | 1,691,428 | 341,785 |
| | Indirect Effects from PCV15 | PCV20 | 2,257,554,999 | 1,691,400 | |
| | | PCV21 | 2,257,814,057 | 1,691,419 | 13,149 |
| | Indirect Effects from PCV20 | PCV21 | 2,258,535,466 | 1,691,384 | |
| | | PCV20 | 2,259,716,210 | 1,691,294 | Dominated |
| 2015-2019 | No indirect Effects | PCV21 | 2,252,167,757 | 1,691,708 | |
| | | PCV20 | 2,259,024,960 | 1,691,325 | Dominated |
| | Indirect Effects from PCV15 | PCV21 | 2,252,531,488 | 1,691,690 | |
| | | PCV20 | 2,260,107,271 | 1,691,272 | Dominated |
| | Indirect Effects from PCV20 | PCV21 | 2,253,285,471 | 1,691,653 | |
| | | PCV20 | 2,262,347,702 | 1,691,168 | Dominated |
| **Societal perspective** | | | | | |
| 2022 | No indirect Effects | PCV20 | 3,073,396,106 | 1,691,426 | |
| | | PCV21 | 3,073,984,363 | 1,691,428 | 298,254 |
| | Indirect Effects from PCV15 | PCV21 | 3,074,312,419 | 1,691,419 | |
| | | PCV20 | 3,074,380,015 | 1,691,400 | Dominated |
| | Indirect Effects from PCV20 | PCV21 | 3,075,620,142 | 1,691,384 | |
| | | PCV20 | 3,078,297,772 | 1,691,294 | Dominated |
| 2015-2019 | No indirect Effects | PCV21 | 3,063,886,165 | 1,691,708 | |
| | | PCV20 | 3,077,078,915 | 1,691,325 | Dominated |
| | Indirect Effects from PCV15 | PCV21 | 3,064,545,499 | 1,691,690 | |
| | | PCV20 | 3,079,040,883 | 1,691,272 | Dominated |
| | Indirect Effects from PCV20 | PCV21 | 3,065,912,251 | 1,691,653 | |
| | | PCV20 | 3,083,102,328 | 1,691,162 | Dominated |

***Table S4**. Mean Costs, QALYs, and ICERs for pneumococcal vaccination of adults aged 50 years with chronic medical conditions (RR = 4), from health system and societal perspectives (per 100,000 population)*

| Serotype distribution data year | Indirect effects assumption | Rest of Canada | | | |
|---|---|---|---|---|---|
| | | Strategies | Costs | QALYs | Sequential ICER |
| **Health system perspective** | | | | | |
| 2022 | No indirect Effects | PCV20 | 464,680,848 | 2,011,123 | |
| | | PCV21 | 465,328,641 | 2,011,124 | 1,225,461 |
| | Indirect Effects from PCV15 | PCV20 | 464,826,930 | 2,011,116 | |
| | | PCV21 | 465,377,339 | 2,011,121 | 101,313 |
| | Indirect Effects from PCV20 | PCV20 | 465,409,270 | 2,011,086 | |
| | | PCV21 | 465,571,537 | 2,011,111 | 6,496 |
| 2015-2019 | No indirect Effects | PCV21 | 463,943,832 | 2,011,196 | |
| | | PCV20 | 465,256,927 | 2,011,093 | Dominated by PCV21 |
| | Indirect Effects from PCV15 | PCV21 | 464,041,335 | 2,011,191 | |
| | | PCV20 | 465,548,697 | 2,011,078 | Dominated by PCV21 |
| | Indirect Effects from PCV20 | PCV21 | 464,234,548 | 2,011,181 | |
| | | PCV20 | 466,153,537 | 2,011,048 | Dominated by PCV21 |
| **Societal perspective** | | | | | |
| 2022 | No indirect Effects | PCV20 | 627,198,060 | 2,011,123 | |
| | | PCV21 | 627,838,260 | 2,011,124 | 1,211,098 |
| | Indirect Effects from PCV15 | PCV20 | 627,450,550 | 2,011,116 | |
| | | PCV21 | 627,922,431 | 2,011,121 | 86,858 |
| | Indirect Effects from PCV20 | PCV21 | 628,258,085 | 2,011,111 | |
| | | PCV20 | 628,457,079 | 2,011,086 | Dominated by PCV21 |
| 2015-2019 | No indirect Effects | PCV21 | 625,416,503 | 2,011,196 | |
| | | PCV20 | 628,203,078 | 2,011,093 | Dominated by PCV21 |
| | Indirect Effects from PCV15 | PCV21 | 625,585,072 | 2,011,191 | |
| | | PCV20 | 628,707,380 | 2,011,078 | Dominated by PCV21 |
| | Indirect Effects from PCV20 | PCV21 | 625,934,531 | 2,011,181 | |
| | | PCV20 | 629,752,807 | 2,011,048 | Dominated by PCV21 |

***Table S9**. Mean Costs, QALYs, and ICERs for pneumococcal vaccination of adults aged 50 years who are unhoused populations (RR = 50), from health system and societal perspectives (per 100,000 population)*

| Serotype distribution data year | Indirect effects assumption | Rest of Canada | | | |
|---|---|---|---|---|---|
| | | Strategies | Costs | QALYs | Sequential ICER |

| | | | | | |
|---|---|---|---|---|---|
| **Health system perspective** | | | | | |
| 2022 | No indirect Effects | PCV20 | 3,357,617,681 | 1,830,102 | |
| | | PCV21 | 3,357,063,583 | 1,830,108 | 74,501 |
| | Indirect Effects from PCV15 | PCV21 | 3,358,625,104 | 1,830,081 | |
| | | PCV20 | 3,359,299,737 | 1,830,021 | Dominated |
| | Indirect Effects from PCV20 | PCV21 | 3,360,857,668 | 1,829,974 | |
| | | PCV20 | 3,365,945,589 | 1,829,704 | Dominated |
| 2015-2019 | No indirect Effects | PCV21 | 3,341,262,194 | 1,830,933 | |
| | | PCV20 | 3,364,334,167 | 1,829,761 | Dominated |
| | Indirect Effects from PCV15 | PCV21 | 3,342,419,325 | 1,830,878 | |
| | | PCV20 | 3,367,650,150 | 1,829,603 | Dominated |
| | Indirect Effects from PCV20 | PCV21 | 3,344,810,135 | 1,830,764 | |
| | | PCV20 | 3,374,449,818 | 1,829,278 | Dominated |
| **Societal perspective** | | | | | |
| 2022 | No indirect Effects | PCV20 | 4,574,652,469 | 1,830,102 | |
| | | PCV21 | 4,575,010,814 | 1,830,108 | 59,872 |
| | Indirect Effects from PCV15 | PCV21 | 4,575,970,943 | 1,830,081 | |
| | | PCV20 | 4,577,528,595 | 1,830,021 | Dominated |
| | Indirect Effects from PCV20 | PCV21 | 4,579,788,418 | 1,829,974 | |
| | | PCV20 | 4,588,892,948 | 1,829,704 | Dominated |
| 2015-2019 | No indirect Effects | PCV21 | 4,546,225,581 | 1,830,933 | |
| | | PCV20 | 4,586,441,928 | 1,829,761 | Dominated |
| | Indirect Effects from PCV15 | PCV21 | 4,548,203,759 | 1,830,878 | |
| | | PCV20 | 4,592,111,670 | 1,829,603 | Dominated |
| | Indirect Effects from PCV20 | PCV21 | 4,552,291,082 | 1,830,764 | |
| | | PCV20 | 4,651,061,119 | 1,827,856 | Dominated |

***Table S10***. *Mean Costs, QALYs, and ICERs for pneumococcal vaccination of adults aged 33 years with immunocompromising conditions (RR = 34), from health system and societal perspectives (per 100,000 population)*

| **Serotype distribution data year** | **Indirect effects assumption** | | **Rest of Canada** | | |
|---|---|---|---|---|---|
| | | **Strategies** | **Costs** | **QALYs** | **Sequential ICER** |
| **Health system perspective** | | | | | |

| | | | | | |
|---|---|---|---|---|---|
| 2022 | No indirect Effects | PCV20 | 1,952,943,411 | 2,349,621 | |
| | | PCV21 | 1,955,332,320 | 2,349,561 | Dominated by PCV20 |
| | Indirect Effects from PCV15 | PCV20 | 1,953,119,872 | 2,349,616 | |
| | | PCV21 | 1,955,390,815 | 2,349,560 | Dominated by PCV20 |
| | Indirect Effects from PCV20 | PCV20 | 1,954,039,509 | 2,349,588 | |
| | | PCV21 | 1,955,696,079 | 2,349,550 | Dominated by PCV20 |
| 2015-2019 | No indirect Effects | PCV21 | 1,953,582,595 | 2,349,616 | |
| | | PCV20 | 1,953,977,126 | 2,349,589 | Dominated by PCV21 |
| | Indirect Effects from PCV15 | PCV21 | 1,953,681,391 | 2,349,613 | |
| | | PCV20 | 1,954,272,626 | 2,349,580 | Dominated by PCV21 |
| | Indirect Effects from PCV20 | PCV21 | 1,954,008,945 | 2,349,603 | |
| | | PCV20 | 1,955,250,791 | 2,349,550 | Dominated by PCV21 |
| **Societal perspective** | | | | | |
| 2022 | No indirect Effects | PCV20 | 2,810,495,976 | 2,349,621 | |
| | | PCV21 | 2,814,712,764 | 2,349,561 | Dominated by PCV20 |
| | Indirect Effects from PCV15 | PCV20 | 2,810,838,311 | 2,349,616 | |
| | | PCV21 | 2,814,826,251 | 2,349,560 | Dominated by PCV20 |
| | Indirect Effects from PCV20 | PCV20 | 2,812,622,461 | 2,349,588 | |
| | | PCV21 | 2,815,418,503 | 2,349,550 | Dominated by PCV20 |
| 2015-2019 | No indirect Effects | PCV21 | 2,811,288,383 | 2,349,616 | |
| | | PCV20 | 2,812,517,337 | 2,349,589 | Dominated by PCV21 |
| | Indirect Effects from PCV15 | PCV21 | 2,811,480,049 | 2,349,613 | |
| | | PCV20 | 2,813,090,629 | 2,349,580 | Dominated by PCV21 |
| | Indirect Effects from PCV20 | PCV21 | 2,812,115,513 | 2,349,603 | |
| | | PCV20 | 2,814,988,400 | 2,349,550 | Dominated by PCV21 |

***Table S11***. *Mean Costs, QALYs, and ICERs for pneumococcal vaccination of adults aged 18-49 years with chronic medical conditions (RR = 4), from health system and societal perspectives (per 100,000 population)*

| **Serotype distribution data year** | **Indirect effects assumption** | **Rest of Canada** | | | |
|---|---|---|---|---|---|
| | | **Strategies** | **Costs** | **QALYs** | **Sequential ICER** |
| **Health system perspective** | | | | | |
| 2022 | No indirect Effects | No vaccine | 352,745,598 | 2,585,328 | |
| | | PCV20 | 352,907,400 | 2,585,406 | 2,084 |
| | | PCV21 | 353,960,132 | 2,585,391 | Dominated by PCV20 |

| | | | | | |
|---|---|---|---|---|---|
| | Indirect Effects from PCV15 | No Vaccine | 352,745,598 | 2,585,328 | |
| | | PCV20 | 352,961,329 | 2,585,405 | 2,830 |
| | | PCV21 | 353,978,069 | 2,585,390 | Dominated by PCV20 |
| | Indirect Effects from PCV20 | No vaccine | 352,745,598 | 2,585,328 | |
| | | PCV20 | 353,242,727 | 2,585,397 | 7,217 |
| | | PCV21 | 354,071,715 | 2,585,388 | Dominated by PCV20 |
| 2015-2019 | No indirect Effects | No vaccine | 352,745,598 | 2,585,328 | |
| | | PCV20 | 353,218,349 | 2,585,398 | 6,820 |
| | | PCV21 | 353,436,472 | 2,585,405 | 31,219 |
| | Indirect Effects from PCV15 | No vaccine | 352,745,598 | 2,585,328 | |
| | | PCV20 | 353,308,840 | 2,585,395 | 8,412 |
| | | PCV21 | 353,466,671 | 2,585,404 | 18,442 |
| | Indirect Effects from PCV20 | No vaccine | 352,745,328 | 2,585,328 | |
| | | PCV21 | 353,566,843 | 2,585,401 | 11,264 |
| | | PCV20 | 353,608,817 | 2,585,388 | Dominated by PCV21 |
| **Societal perspective** | | | | | |
| 2022 | No indirect Effects | No vaccine | 494,791,536 | 2,585,328 | |
| | | PCV20 | 500,772,943 | 2,585,406 | 77,056 |
| | | PCV21 | 502,283,380 | 2,585,391 | Dominated by PCV20 |
| | Indirect Effects from PCV15 | No vaccine | 494,791,536 | 2,585,328 | |
| | | PCV20 | 500,869,469 | 2,585,405 | 79,743 |
| | | PCV21 | 502,315,486 | 2,585,390 | Dominated by PCV20 |
| | Indirect Effects from PCV20 | No vaccine | 494,791,536 | 2,585,328 | |
| | | PCV20 | 501,373,148 | 2,585,397 | 95,544 |
| | | PCV21 | 502,483,107 | 2,585,388 | Dominated by PCV20 |
| 2015-2019 | No indirect Effects | No vaccine | 494,791,536 | 2,585,328 | |
| | | PCV21 | 501,341,472 | 2,585,405 | 85,840 |
| | | PCV20 | 501,332,149 | 2,585,398 | Extendedly dominated |
| | Indirect Effects from PCV15 | No vaccine | 494,791,536 | 2,585,328 | |
| | | PCV21 | 501,395,525 | 2,585,404 | 87,450 |
| | | PCV20 | 501,494,121 | 2,585,395 | Dominated by PCV21 |
| | Indirect Effects from PCV20 | No vaccine | 494,791,536 | 2,585,328 | |
| | | PCV21 | 501,574,824 | 2,585,401 | 93,040 |
| | | PCV20 | 502,031,063 | 2,585,388 | Dominated by PCV21 |

**Table S12**. Mean Costs, QALYs, and ICERs for pneumococcal vaccination of adults aged 18-49 years of unhoused population (RR = 50), from health system and societal perspectives (per 100,000 population)

| Serotype distribution data year | Indirect effects assumption | Rest of Canada | | | |
|---|---|---|---|---|---|
| | | Strategies | Costs | QALYs | Sequential ICER |
| **Health system perspective** | | | | | |
| 2022 | No indirect Effects | PCV20 | 2,841,894,983 | 2,459,641 | |
| | | No vaccine | 2,873,179,021 | 2,458,780 | Dominated by PCV21 |
| | | PCV21 | 2,849,561,000 | 2,459,460 | Dominated by PCV20 |
| | Indirect Effects from PCV15 | PCV20 | 2,842,566,191 | 2,459,625 | |
| | | No vaccine | 2,873,179,021 | 2,458,780 | Dominated by PCV21 |
| | | PCV21 | 2,849,776,632 | 2,459,455 | Dominated by PCV20 |
| | Indirect Effects from PCV20 | PCV20 | 2,846,023,666 | 2,459,541 | |
| | | No vaccine | 2,873,179,021 | 2,458,780 | Dominated by PCV21 |
| | | PCV21 | 2,850,897,847 | 2,459,428 | Dominated by PCV20 |
| 2015-2019 | No indirect Effects | PCV21 | 2,842,942,021 | 2,459,625 | |
| | | No vaccine | 2,873,179,021 | 2,458,780 | Dominated by PCV20 |
| | | PCV20 | 2,845,853,567 | 2,459,542 | Dominated by PCV21 |
| | Indirect Effects from PCV15 | PCV21 | 2,843,317,086 | 2,459,616 | |
| | | No vaccine | 2,873,179,021 | 2,458,780 | Dominated by PCV20 |
| | | PCV20 | 2,846,956,209 | 2,459,516 | Dominated by PCV21 |
| | Indirect Effects from PCV20 | PCV21 | 2,844,554,773 | 2,459,586 | |
| | | No vaccine | 2,873,179,021 | 2,458,780 | Dominated by PCV20 |
| | | PCV20 | 2,850,559,449 | 2,459,428 | Dominated by PCV21 |
| **Societal perspective** | | | | | |
| 2022 | No indirect Effects | PCV20 | 4,085,198,206 | 2,459,641 | |
| | | No vaccine | 4,134,168,120 | 2,458,780 | Dominated by PCV21 |
| | | PCV21 | 4,098,316,594 | 2,459,460 | Dominated by PCV20 |
| | Indirect Effects from PCV15 | PCV20 | 4,086,369,518 | 2,459,625 | |
| | | No vaccine | 4,134,168,120 | 2,458,780 | Dominated by PCV21 |
| | | PCV21 | 4,098,692,974 | 2,459,455 | Dominated by PCV20 |
| | Indirect Effects from PCV20 | PCV20 | 4,092,403,889 | 2,459,541 | |
| | | No vaccine | 4,134,168,120 | 2,458,780 | Dominated by PCV21 |
| | | PCV21 | 4,100,650,063 | 2,459,428 | Dominated by PCV20 |
| | No indirect Effects | PCV21 | 4,086,724,549 | 2,459,625 | |

| | | | | | |
|---|---|---|---|---|---|
| 2015-2019 | | No vaccine | 4,134,168,120 | 2,458,780 | Dominated by PCV20 |
| | | PCV20 | 4,092,140,500 | 2,459,542 | Dominated by PCV21 |
| | Indirect Effects from PCV15 | PCV21 | 4,087,379,064 | 2,459,616 | |
| | | No vaccine | 4,134,168,120 | 2,458,780 | Dominated by PCV20 |
| | | PCV20 | 4,094,065,015 | 2,459,516 | Dominated by PCV21 |
| | Indirect Effects from PCV20 | PCV21 | 4,089,539,079 | 2,459,586 | |
| | | No vaccine | 4,134,168,120 | 2,458,780 | Dominated by PCV20 |
| | | PCV20 | 4,100,354,503 | 2,459,428 | Dominated by PCV21 |
